# *In vivo* coupling of dendritic complexity with presynaptic density in primary tauopathies

**DOI:** 10.1101/2020.12.24.20248838

**Authors:** Elijah Mak, Negin Holland, P. Simon Jones, George Savulich, Audrey Low, Maura Malpetti, Sanne S Kaalund, Luca Passamonti, Timothy Rittman, Rafael Romero-Garcia, Roido Manavaki, Guy B. Williams, Young T. Hong, Tim D. Fryer, Franklin I. Aigbirhio, John T O’Brien, James B Rowe

**Author notes:** **Corresponding Author:** Dr Negin Holland MRCP, BSc, Association of British Neurologists Clinical Research Fellow, University of Cambridge, Herschel Smith Building, Robinson Way, Cambridge, CB2 0SZ, UK. Joint first authors (Elijah Mak, Negin Holland). Joint senior authors (John T O’Brien, James B Rowe).

## Abstract

Understanding the cellular underpinnings of neurodegeneration remains a challenge; loss of synapses and dendritic arborisation are characteristic and can be quantified i*n vivo*, with [^11^C]UCB-J PET and MRI-based Orientation Dispersion Imaging (ODI), respectively. We aimed to assess how both measures are correlated, in 4R-tauopathies of Progressive Supranuclear Palsy (PSP-RS; n = 22) and amyloid-negative (determined by [^11^C]PiB PET) Corticobasal Degeneration (CBD; n =14), as neurodegenerative disease models, in this proof-of-concept study. Compared to controls (n = 27), PSP-RS and CBD patients had widespread reductions in cortical ODI, and [^11^C]UCB-J non-displaceable binding potential (BP_ND_) in excess of atrophy. In PSP-RS and CBD separately, regional cortical ODI was significantly associated with [^11^C]UCB-J BP_ND_ in disease-associated regions (p < 0.05, FDR corrected). Our findings indicate that reductions in synaptic density and dendritic complexity in PSP-RS and CBD are more severe and extensive than atrophy. Furthermore, both measures are tightly coupled *in vivo*, furthering our understanding of the pathophysiology of neurodegeneration, and applicable to studies of early neurodegeneration with a safe and widely available MRI platform.

## 1. INTRODUCTION

Abnormal dendritic morphology, and synaptic pathology are increasingly recognised as hallmarks of neurodegeneration, prior to cell death and atrophy, and correlate closely with cognitive dysfunction (Clare et al., 2010; DeKosky and Scheff, 1990; Herms and Dorostkar, 2016; Terry et al., 1991). However, quantification of synaptic pathology *in vivo* has until recently posed a challenge. Much of our knowledge about synaptic loss in neurodegeneration stems from animal and *post mortem* studies. For example, with frontotemporal lobar degeneration pathology (Coyle-Gilchrist et al., 2016) including Frontotemporal lobe Dementia (FTD), Progressive Supranuclear Palsy (PSP) and Corticobasal Degeneration (CBD), there is 30-50% loss of synapses, as determined by *post mortem* synaptophysin assays (Bigio et al., 2001; Lipton et al., 2001).

To quantify this pathophysiological change *in vivo* and uncover the early stages of neurodegeneration, would enable better design of clinical trials, accurate measurement of therapeutic response, and early intervention. *In vivo* cerebral glucose metabolism measured with [^18^F]FDG Positron Emission Tomography (PET), has often been interpreted as a synaptic density marker. Reductions are clinically relevant, for example, there are hippocampal reductions of [^18^F]FDG uptake in mild cognitive impairment (Mosconi et al., 2005) and reduced frontotemporal uptake in frontotemporal lobar degeneration (Blin et al., 1992; Juh et al., 2004); however, [^18^F]FDG is an indirect marker. In contrast, the recently, developed radioligand [^11^C]UCB-J ((*R*)-1-((3-(methyl-11C)pyridin-4-yl)methyl)-4-(3,4,5-trifluorophenyl)pyr-rolidin-2-one), allows direct quantification of synaptic density based on its affinity for the ubiquitously expressed presynaptic vesicle glycoprotein SV2A (Finnema et al., 2017, 2016). This ligand shows up-to 40% loss of hippocampal synapses in patients with Alzheimer’s disease (Chen et al., 2018), and a 20-50% global reduction in PSP-RS/CBD (Holland et al., 2020) correlating with cognitive dysfunction. However, PET has limited scalability in diagnostics and trials, because of cost, specialist facilities required, and radiation limitations on repeated assessment.

Here we propose that with the loss of synapses, pre-synaptic vesicle density would correlate with changes in post-synaptic dendritic complexity. While diffusion tensor imaging (DTI) has proved to be a valuable non-invasive technique to probe microstructural white matter integrity, it has limited utility to characterise such changes in the cortex due to the isotropic water diffusion of the grey matter. Therefore, there is a need for biomarkers capable of evaluating functionally relevant microstructure noninvasively. To address the limitations of DTI, Neurite Orientation Dispersion Imaging (NODDI) is a multi-compartment biophysical diffusion imaging model that is capable of disentangling the diffusion signal arising from distinct tissue compartments, such as extracellular, intracellular water and cerebrospinal fluid (Zhang et al., 2012). This multi-compartmentalisation of the diffusion signal enables metrics such as Neurite Density Index (NDI) and Orientation Dispersion Index (ODI) to characterise the amount of neurites as well as the variability of neurite orientations respectively. Both the clinical feasibility and relevance of NODDI alterations have been successfully demonstrated in normal ageing and dementia (Cox et al., 2016; Slattery et al., 2017, 2015; Zhang et al., 2012). However, the biological substrates of NODDI are still unclear due to limited evidence of cross-validation among NODDI metrics and alternative proxies of grey matter microstructure. Recent studies have shown that NODDI Orientation Dispersion Index (ODI) is correlated with histologically-derived indices of neurite dispersion (Grussu et al., 2017; Schilling et al., 2018), highlighting its potential viability as a proxy of underlying biological changes in grey matter microstructure. Of particular relevance to our current study, reductions in cortical ODI has been found in a tau-transgenic mice model, entirely consistent with the severe extent of dendritic degeneration (Colgan et al., 2016). To the best of our knowledge, there has not been any attempt to characterise the coupling of cortical ODI with measurements of synaptic pathology (with [^11^C]UCB-J PET imaging). Compared to PET imaging, the ODI from NODDI would have the advantage of wide scalability in multicentre studies, repeatable assays without ionising radiation, and lower cost within a clinically feasible acquisition time (∼ 15 minutes).

In this proof-of-concept study, we focus on two rapidly progressive neurodegenerative tauopathies, progressive supranuclear palsy – Richardson’s Syndrome (PSP-RS) and corticobasal degeneration (CBD) as disease models where grey matter atrophy although present, is not extensive unlike in Alzheimer’s disease where we would expect generalised atrophy; this allows us to examine cortical microstructure without the influence of atrophy. Pathologically, both PSP-RS and CBD are associated with accumulation of both cortical and subcortical 4-repeat tau (Dickson et al., 2011; Kovacs et al., 2020; Rösler et al., 2019; Schofield et al., 2012), and clinically present with a movement disorder (Armstrong et al., 2013; Höglinger et al., 2017; Litvan et al., 1996) and cognitive decline (Burrell et al., 2014). Both conditions are associated with cortical and subcortical atrophy (Jabbari et al., 2019) but we have previously shown significant widespread presynaptic loss extending beyond atrophic areas (Holland et al., 2020) concordant with findings from studies utilising electromagnetoencephalography where impairment in cortical physiology is seen in both atrophic and minimally atrophic regions (Cope et al., 2018; Hughes et al., 2014; Sami et al., 2018). Having illustrated presynaptic loss as a potential explanation for impairment in cortical physiology, we aimed to investigate the synaptic bouton at large by also focusing on the post-synaptic compartment, by utilising a non-invasive MRI method such as ODI, as an index of dendritic complexity.

There is a very high clinicopathological correlation between 4R-tauopathy and the classical presentation of PSP-RS-Richardson’s syndrome. The clinical spectrum of CBD is diverse; the pathology of CBD is a major cause of the corticobasal syndrome (CBS), which is often mimicked by Alzheimer’s disease or sometimes PSP-RS pathology (Alexander et al., 2014). For this reason, CBD generally refers to the pathology and CBS to the clinical syndrome. This strategy reduces the likelihood of mixing amyloid pathology from Alzheimer’s disease with the primary 4R-tauopathy of PSP-RS and CBD. We therefore infer that patients with CBS in whom Alzheimer’s disease is excluded (with [^11^C]PiB PET) have probable CBD, but acknowledge that other pathologies are also likely.

We test the hypothesis that the functionally relevant loss of pre-synaptic density caused by PSP-RS and CBD is correlated to the changes in post-synaptic dendritic complexity, independent of changes in grey matter atrophy.

## 2. MATERIALS AND METHODS

### 2.1. Experimental design

Patients were recruited from the Cambridge tertiary neurology clinic with for PSP-RS and CBS/CBD. Healthy volunteers were recruited from the UK National Institute for Health Research Join Dementia Research (JDR) register. Patients had either probable PSP-RS– Richardson Syndrome (Höglinger et al., 2017), or CBS with possible/probable CBD (Armstrong et al., 2013). Participants underwent a clinical and cognitive assessment including measures of disease severity **(Table 1)**, 3T MRI, and [^11^C]UCB-J PET. Patients with CBD underwent amyloid PET imaging using Pittsburgh Compound B ([^11^C]-PiB). Only those with a negative amyloid status are included in the subsequent analysis, as determined by a [^11^C]-PiB standardised uptake value ratio (SUVR; 50-70 minutes post injection; whole cerebellum reference tissue) less than 1.21 (Centiloid scale 19 (Jack et al., 2017)); on this basis 12 out of 26 CBS patients were excluded. The research protocol was approved by the Cambridge Research Ethics Committee and the Administration of Radioactive Substances Advisory Committee. All participants provided written informed consent in accordance with the Declaration of Helsinki.

**Table 1.**
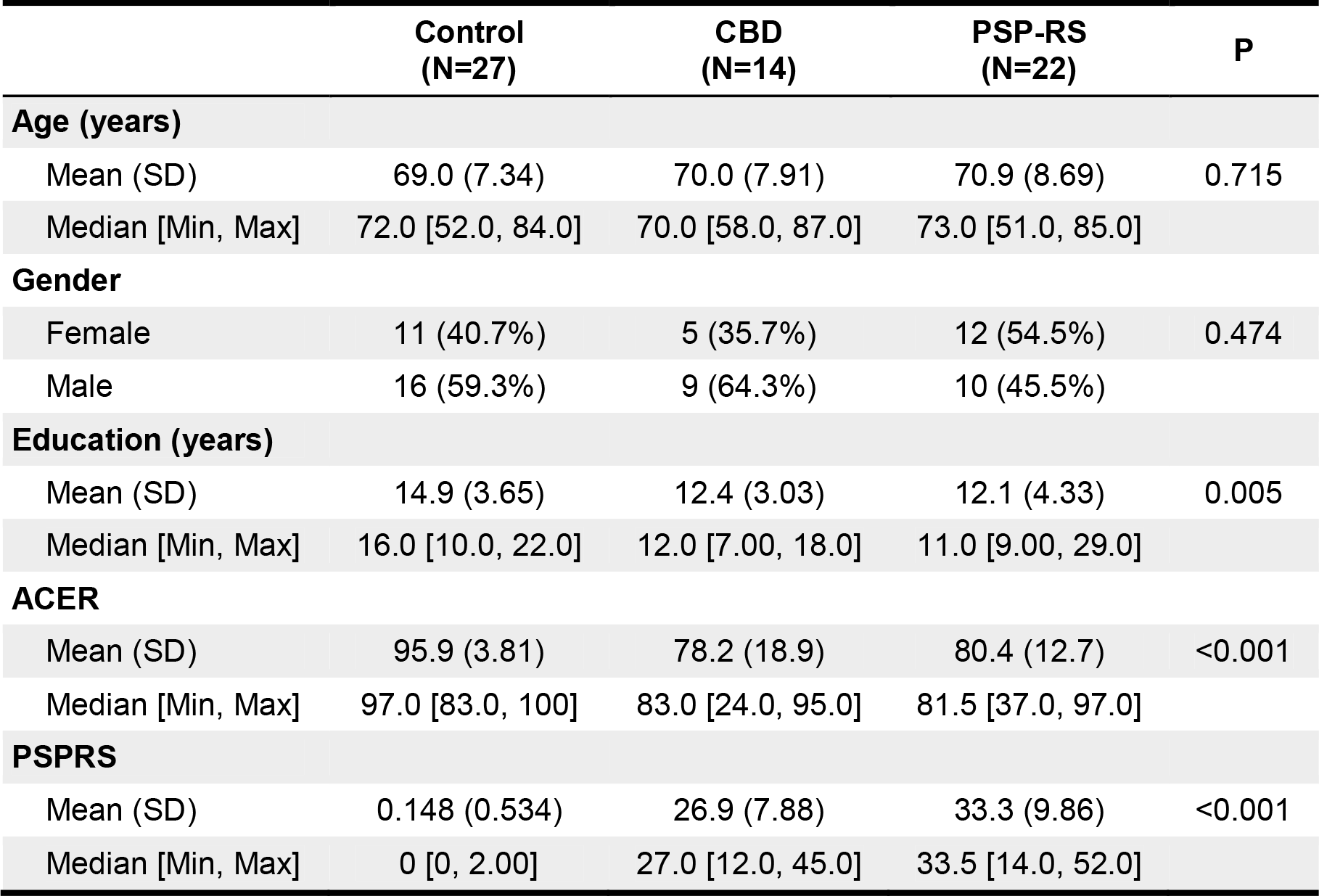
Demographics and clinical variables. Abbreviations: CBD = Corticobasal Degeneration defined as amyloid negative corticobasal syndrome, PSP-RS = Progressive Supranuclear Palsy - Richardson’s syndrome, PSPRS = Progressive Supranuclear Palsy Rating Scale; ACER = Addenbrooke’s Cognitive Examination-Revised. Parameters are expressed as mean ± standard deviation (SD). P values are shown for analysis of variance across groups or chi-squared test as appropriate.

|  | Control<br>(N=27) | CBD<br>(N=14) | PSP-RS<br>(N=22) | P |
| --- | --- | --- | --- | --- |
| **Age (years)** |  |  |  |  |
| Mean (SD) | 69.0 (7.34) | 70.0 (7.91) | 70.9 (8.69) | 0.715 |
| Median [Min, Max] | 72.0 [52.0, 84.0] | 70.0 [58.0, 87.0] | 73.0 [51.0, 85.0] |  |
| **Gender** |  |  |  |  |
| Female | 11 (40.7%) | 5 (35.7%) | 12 (54.5%) | 0.474 |
| Male | 16 (59.3%) | 9 (64.3%) | 10 (45.5%) |  |
| **Education (years)** |  |  |  |  |
| Mean (SD) | 14.9 (3.65) | 12.4 (3.03) | 12.1 (4.33) | 0.005 |
| Median [Min, Max] | 16.0 [10.0, 22.0] | 12.0 [7.00, 18.0] | 11.0 [9.00, 29.0] |  |
| **ACER** |  |  |  |  |
| Mean (SD) | 95.9 (3.81) | 78.2 (18.9) | 80.4 (12.7) | <0.001 |
| Median [Min, Max] | 97.0 [83.0, 100] | 83.0 [24.0, 95.0] | 81.5 [37.0, 97.0] |  |
| **PSPRS** |  |  |  |  |
| Mean (SD) | 0.148 (0.534) | 26.9 (7.88) | 33.3 (9.86) | <0.001 |
| Median [Min, Max] | 0 [0, 2.00] | 27.0 [12.0, 45.0] | 33.5 [14.0, 52.0] |  |

### 2.2. Neuroimaging processing

#### 2.2.1. T1-weighted MRI

T1-MPRAGE was acquired on the Siemens Magnetom 3T PRISMA scanner (TE = 2.93 ms, TR = 2s, slice thickness = 1.1 mm, resolution = 1.1 mm^3^ isotropic, 208 slices). The data were processed using Computational Anatomy Toolbox in SPM12. The T1-MPRAGE images were then segmented into grey matter, white matter, and CSF images by using a unified tissue segmentation technique after image intensity non-uniformity correction was performed. Regional cortical thickness was derived in CAT12 based on the projection-based thickness method (Dahnke et al., 2013), which uses topology correction. (Yotter et al., 2011a) and spherical mapping (Yotter et al., 2011b). Previous studies have shown that CAT12 produce reliable estimates of cortical thickness, yielding larger effect sizes in case-control comparisons (Seiger et al., 2018). All segmentations were visually inspected by 3 authors (E.M, A.L. and M.M). 3 PSP-RS subjects were excluded due to sub-optimal contrast between grey and white matter tissue, resulting in unsatisfactory segmentations.

#### 2.2.2. Diffusion-weighted MRI

Diffusion scans were acquired on Siemens Magnetom Prisma scanner (TE = 75.6ms, TR = 2.4s, slice thickness = 1.75mm, 98 directions, 104 slices, bvals = 300, 1000, 2000). Diffusion datasets were pre-processed with FSL-FDT (FMRIB’s Diffusion Toolbox). Firstly, the DWI data were stripped of nonbrain tissue using the Brain Extraction Tool. The resulting brain masks were visually inspected for anatomic fidelity. Eddy currents and head movements were corrected with “eddy” in FSL (Version 6.0.1). TOPUP was applied to correct for estimating and correcting susceptibility induced distortions. The b0 volume from the reversed phase-encode blip was used in TOPUP for the estimation and correction of susceptibility induced distortion. Quantitative identification of slices with signal loss was performed in “eddy” and these volumes were replaced by non-parametric predictions using the Gaussian process (Andersson et al., 2016).The b-matrix was subsequently reoriented by applying the rotational part of the affine transformation used during eddy correction (Leemans and Jones, 2009). Next, ODI maps were derived from the eddy-corrected datasets using the Microstructural Diffusion Toolbox (MDT) (Harms et al., 2017). There has been recent debate over the validity of NODDI’s assumption of a fixed intrinsic diffusivity across the brain, especially in the grey matter. Therefore, to optimise analyses of ODI, we set the intrinsic diffusivity to 1.1 (0.1) x 10^−3^ mm^2^/s as previously proposed (Fukutomi et al., 2018; Genç et al., 2018).

#### 2.2.3. Positron Emission Tomography (PET)

A subset of patients underwent dynamic PET imaging (Controls = 19, PSP-RS = 20, CBD = 12) on a GE SIGNA PET/MR (GE Healthcare, Waukesha, USA) for 90 minutes starting immediately after [^11^C]UCB-J injection (median injected activity: 351 ± 107 MBq, injected mass ≤ 10 μg), with attenuation correction including the use of a multi-subject atlas method (Burgos et al., 2014; Prados et al., 2016) and also improvements to the MRI brain coil component (Manavaki et al., 2019). Each emission image series was aligned using SPM12 (www.fil.ion.ucl.ac.uk/spm/software/spm12/) to ameliorate the effect of patient motion, then rigidly registered to a T1-weighted MRI acquired during PET data. To quantify SV2A density, parametric maps of [^11^C]UCB-J non-displaceable binding potential (BP_ND_) was determined using a basis function implementation of the simplified reference tissue model (Wu and Carson, 2002), with the reference tissue defined in the centrum semiovale (Koole et al., 2019; Rossano et al., 2019).

#### 2.2.4. Extraction of regional measurements from T1, ODI and [^11^C]UCB-J BP_ND_

ODI and [^11^C]UCB-J maps were coregistered to the skull-stripped T1 brain image using rigid registrations in Advance Normalisation Tools (ANTS; http://stnava.github.io/ANTs/). The accuracy of all coregistrations was visually inspected. The coregistered ODI and [^11^C]UCB-J volumes were projected to the surface for cortical analyses. To that end, we applied a weighted-mean method that uses a Gaussian kernel for mapping along the normal, based on the recommended settings in CAT12. Finally, ROI extraction was performed within the surface space using the Desikan Killiany atlas. We used a modified Hammers atlas to extract measurements from the subcortical regions and midbrain. The atlas was subsequently transposed into the native spaces of ODI and [^11^C]UCB-J respectively using the inverse of transformations from the coregistrations between ODI or [^11^C]UCB-J and T1 MPRAGE. Mean regional ODI, [^11^C]UCB-J and grey matter volumes were extracted using *fslstats* in FSL. Finally, each subject has the following imaging measurements: regional cortical thickness, cortical ODI, cortical [^11^C]UCB-J BP_ND_, as well as subcortical and midbrain grey matter volumes, ODI and [^11^C]UCB-J BP_ND._

### 2.3. Statistical Analysis

We used R to compare demographic variables between the diagnostic groups using ANOVA, Kruskal Wallis and chi-square tests where appropriate. To investigate regional group differences of grey matter, ODI and [^11^C]UCB-J BP_ND,_ we used non-parametric permutation-based inference, implemented using Permutation Analysis of Linear Models (PALM; https://fsl.fmrib.ox.ac.uk/fsl/fslwiki/PALM) in MATLAB, adjusted for age (5000 permutations) (Winkler et al., 2014). A key advantage of permutation-based approach is the robustness of statistics to heteroscedasticity and its minimal assumptions on data distributions. Next, non-parametric permutation tests for Biological Parametric Mapping (BPM) were used to assess whole-brain inter-regional associations between ODI and [^11^C]UCB-J BP_ND_ across the patient sample, as well as within CBS and PSP-RS groups separately. Correlational analyses were adjusted for age and local grey matter thickness / volume. Statistical results were adjusted using the False Discovery Rate (FDR) correction across the cortical, subcortical regions and the midbrain (81 ROIs). Mean values, standard deviations, P values and Cohen’s D were overlaid on 2D brain templates using the *ggseg* package in R (Mowinckel and Vidal-Piñeiro, 2019). The derived data that support the findings of this study are available from the corresponding author, upon reasonable request for academic (non-commercial) purposes.

## 3. RESULTS

### 3.1. Demographics

Clinical and demographic information are summarised in **Table 1**. Patients and controls were matched in age and gender although education years were significantly lower in PSP-RS relative to controls (Post-hoc Dunn’s Test after Kruskal-Wallis test; p < 0.001). Patients with PSP-RS and CBD had lower total ACE-R scores compared to controls (p < 0.001).

### 3.2. Group comparisons of grey matter atrophy, ODI and [^11^C]UCB-J BP_ND_

For all groups, mean ± standard deviation (SD) distributions of regional cortical thickness, subcortical grey matter volumes, ODI and [^11^C]UCB-J BP_ND_ are overlaid on brain templates in **Figure 1** and **Figure 2**. Permutation-based statistical comparisons across all imaging measurements are reported for PSP-RS and CBD relative to controls, adjusted for age and FDR corrected in **Figure 3** and **Figure 4**.

**Figure 1.**
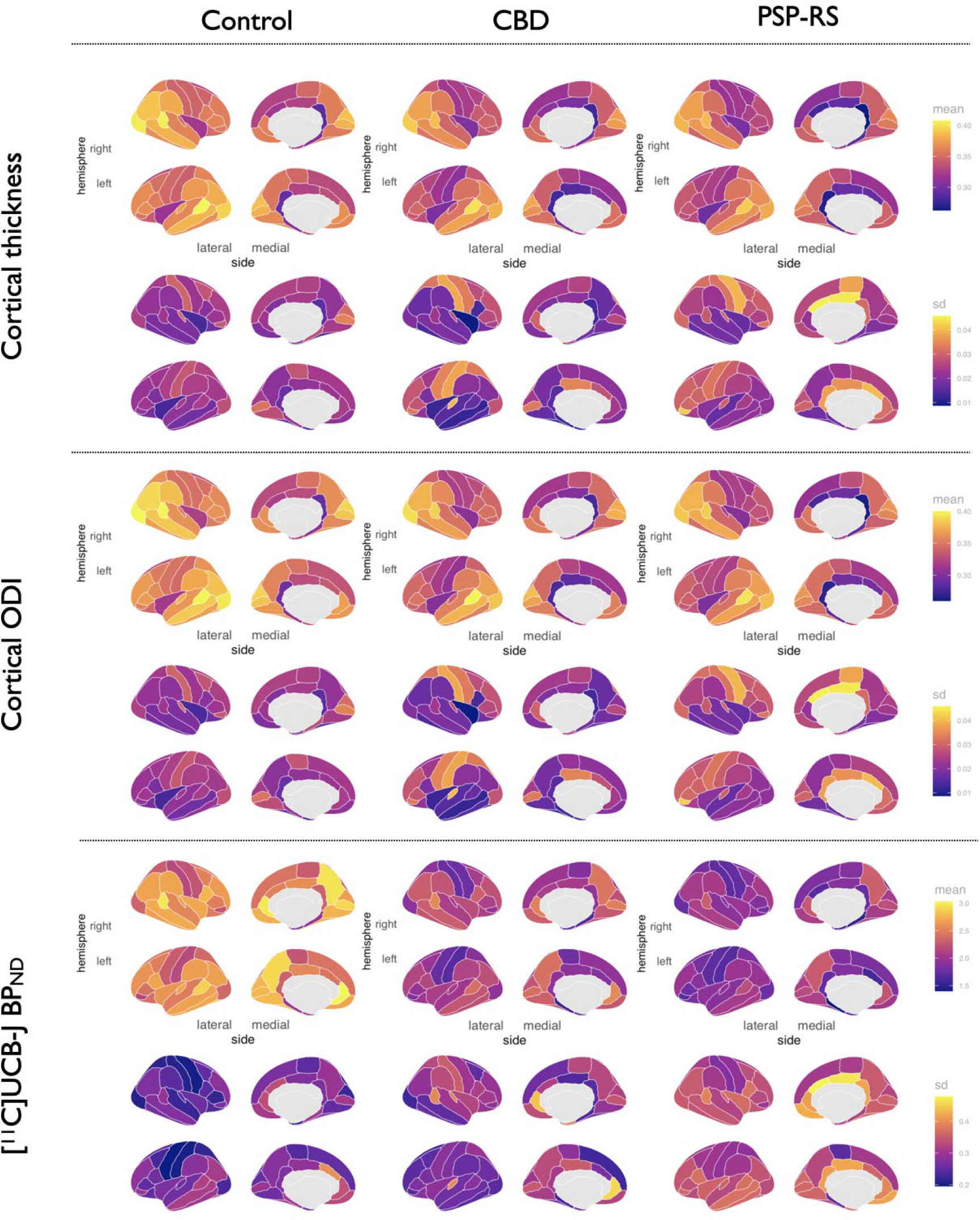
Regional distributions of cortical thickness, cortical ODI and cortical [^11^C]-UCB-J BP_ND_ across controls, CBD, and PSP-RS. Abbreviations: ODI = Orientation Dispersion Index, CBD = amyloid negative Corticobasal Syndrome; PSP-RS = Progressive Supranuclear Palsy-Richardson’s Syndrome; sd = Standard deviation.

**Figure 2.**
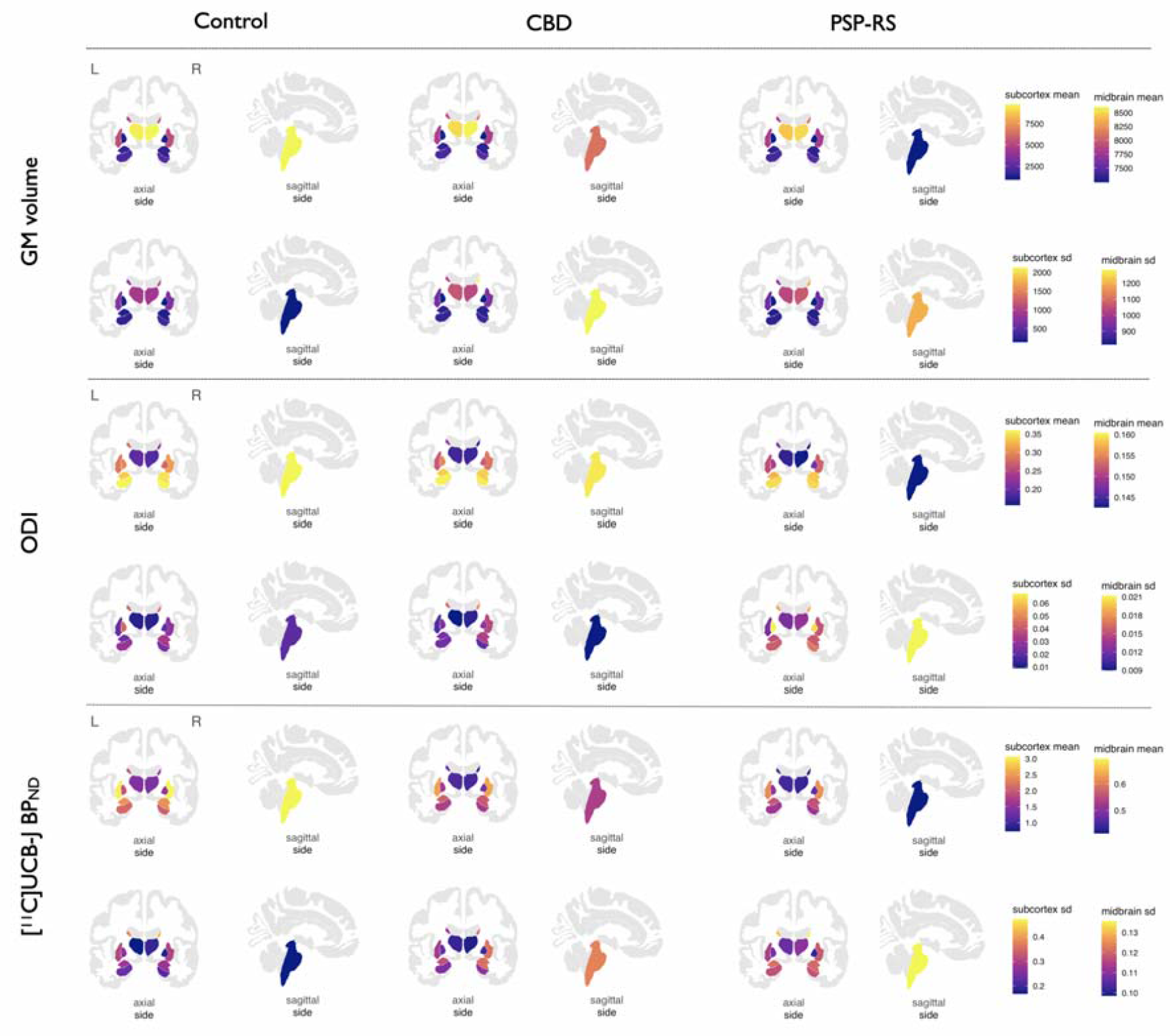
Subcortical distributions of grey matter volumes, ODI and [^11^C]-UCB-J BP_ND_ across controls, CBD, and PSP-RS. Abbreviations: ODI = Orientation Dispersion Index, CBD = amyloid negative Corticobasal Syndrome; PSP-RS = Progressive Supranuclear Palsy-Richardson’s Syndrome; sd = standard deviation; GM = grey matter. For ease of visualisation the colours in the brainstem represent the midbrain only.

**Figure 3.**
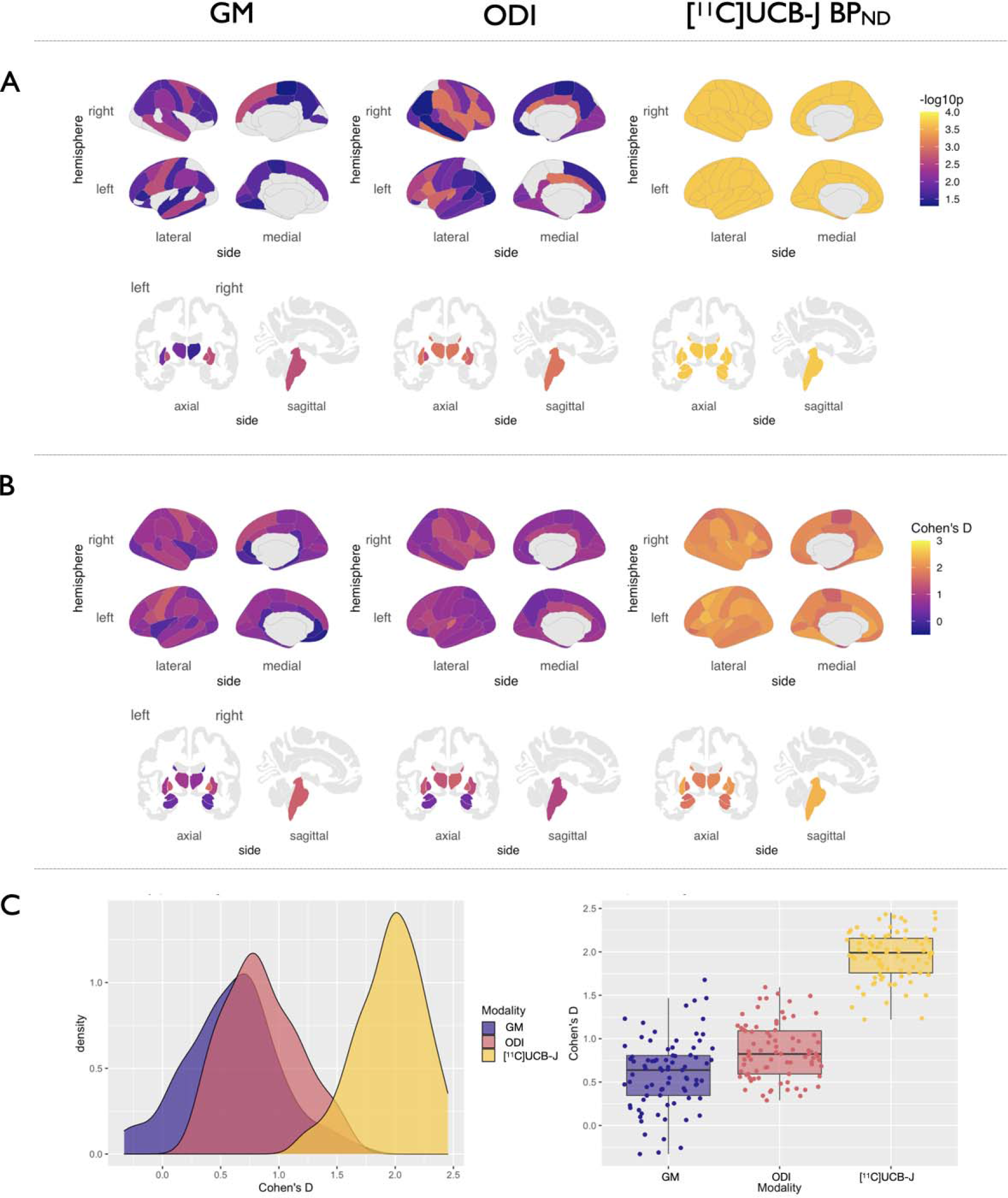
GM atrophy, reduced ODI and [^11^C]UCB-J BP_ND_ in PSP-RS vs Controls. **A:** P-values of regions surviving FDR correction are visualised on cortical and subcortical brain templates. **B:** Density plots of distributions of effect sizes. **C:** Boxplots of effect sizes of grey matter atrophy, ODI and [^11^C]UCB-J BP_ND_ from the group comparisons. Abbreviations: ODI = Orientation Dispersion Index; PSP-RS = Progressive Supranuclear Palsy-Richardson’s Syndrome; FDR = False Discovery Rate; GM = grey matter.

**Figure 4.**
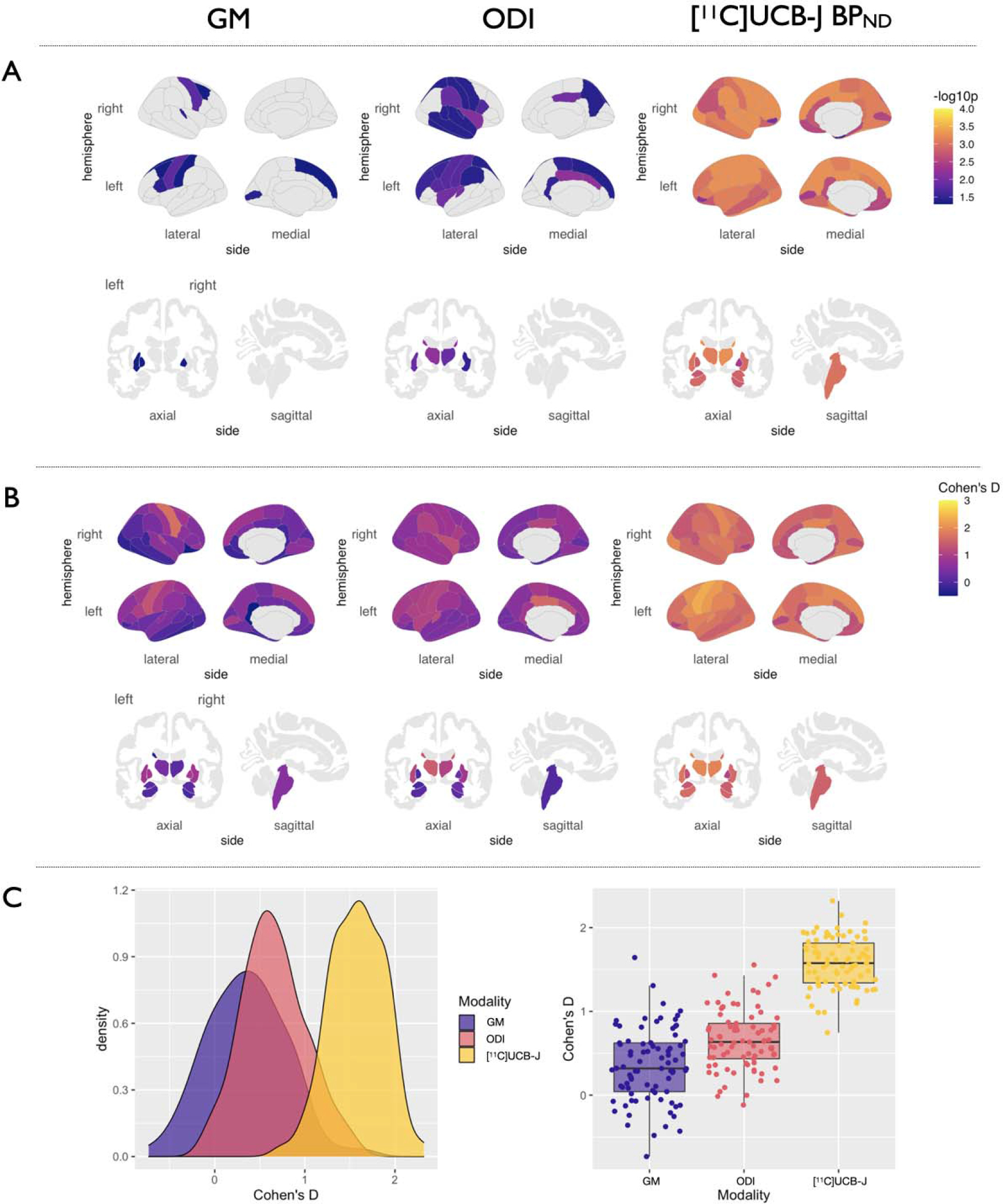
GM atrophy, reduced ODI and [^11^C]UCB-J BP_ND_ in CBD vs Controls. **A:** P-values of regions surviving FDR correction are visualised on cortical and subcortical brain templates. **B:** Density plots of distributions of effect sizes. **C:** Boxplots of effect sizes of grey matter atrophy, ODI and [^11^C]UCB-J BP_ND_ from the group comparisons. Abbreviations: ODI = Orientation Dispersion Index; FDR = False Discovery Rate; GM = grey matter; CBD = amyloid negative Corticobasal Syndrome.

#### 3.2.1. Progressive supranuclear palsy – Richardson’s Syndrome

Statistical results and corresponding effect sizes from the comparisons between PSP-RS and controls illustrated in **Figure 3A-C**. Non-parametric permutation-based tests showed that, relative to controls, the PSP-RS group exhibited significant cortical thinning particularly in the motor cortex and frontal cortices; subcortical atrophy was found in the thalamus, putamen and pallidum and midbrain. In PSP-RS, significant reductions in cortical ODI were more widespread than areas affected by atrophy, including multiple areas within the cortical mantle and also subcortically including the basal ganglia. Post-hoc paired T-Tests of the regional Fisher’s transformed Cohen’s D indicated that the regional effect sizes were significantly larger for ODI compared to grey matter atrophy **(Figure 3C)**. As expected, the PSP-RS group showed extensive and severe reductions of [^11^C]UCB-J BP_ND_ across the cortex and subcortical regions compared to controls, replicating our previous findings in a smaller sample (Holland et al., 2020). Across the whole brain, reductions in [^11^C]UCB-J BP_ND_ yielded the largest effect size relative to both ODI and grey matter atrophy.

#### 3.2.2. Corticobasal Degeneration

Statistical results and corresponding effect sizes from the comparisons between CBD and controls are illustrated in **Figure 4A-C**. Relative to controls, the CBD group showed focal cortical thinning in motor cortex, superior frontal cortex and the occipital lobe, as well as atrophy in the left putamen and bilateral pallidum. In contrast, widespread ODI reductions were found extending beyond the atrophy-affected motor cortices to other regions that were relatively preserved from atrophy, notably the temporo-parietal and cingulate cortices. In addition, there was bilateral significant ODI reductions in the caudate and putamen, both of which showed no significant grey matter atrophy. Accordingly, regional Cohen’s D of ODI was significantly larger than that of grey matter atrophy **(Figure 4C)**. Finally, [^11^C]UCB-J BP_ND_ comparisons revealed a generalised and widespread extent of significantly reduced [^11^C]UCB-J BP_ND_ across the cortex and subcortical regions compared to controls.

### 3.3. Regional associations of ODI with [^11^C]UCB-J BP_ND_

Non-parametric permutation models were used to assess the inter-regional associations between ODI and [^11^C]UCB-J BP_ND_ across the full patient sample, while sensitivity analyses were conducted for PSP-RS and CBD groups separately. To visualise the spatial patterns of these regional correlations, accounting for age and regional grey matter atrophy, we projected the p values on brain templates **(Figure 5)**. Within the total sample of CBD and PSP-RS patients, multiple cortical and subcortical regions demonstrated local positive associations where [^11^C]UCB-J BP_ND_ locally predicted ODI, i.e. across subjects, both presynaptic density and dendritic arborisation showed reciprocal associations within the same region **(Figure 5A)**. Scatter plots for each significant local association are shown in **Supplementary Figure 1**. The regions showing the strongest correlations between ODI and [^11^C]UCB-J BP_ND_ were primarily in bilateral pre and postcentral gyri and the prefrontal cortex, but also extended to subcortical areas including the basal ganglia, thalamus and the midbrain. When the analyses were restricted to the CBD sample, the spatial extent of local associations was markedly attenuated, although significant associations were still present within the bilateral pre and post central gyri, and isolated regions in the frontal and occipital lobe **(Figure 5B)**. Within the PSP-RS group, [^11^C]UCB-J BP_ND_ was significantly correlated with ODI in a widespread spatial pattern similar to that of the total sample. Peak correlations were identified within the motor cortex and cingulate regions **(Figure 5C)**, but also included the thalami, midbrain and parts of the basal ganglia. These analyses did not include the control group and is thus not indicative of a group effect. Nevertheless, to determine the specificity of the coupling between ODI and [^11^C]UCB-J BP_ND,_ we ran the same non-parametric permutation model on the controls, accounting for age and regional grey matter atrophy. This analysis did not yield any significant local associations that retained statistical significance after FDR correction.

**Figure 5.**
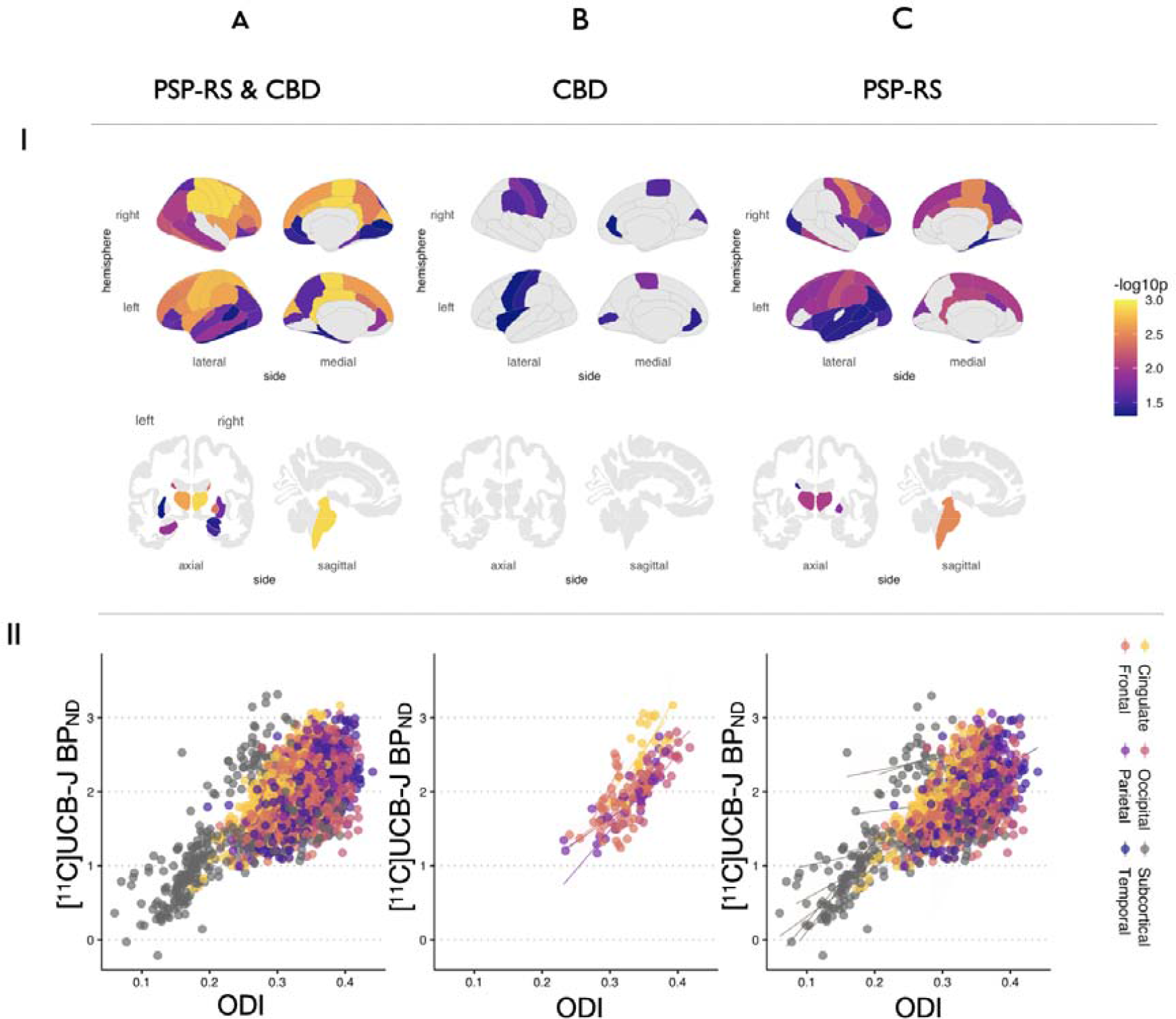
Local associations between ODI and [^11^C]UCB-J BP_ND_ in (A) total sample of patients, (B) CBD and (C) PSP-RS separately. I: Statistical p values for regions surviving FDR correction are overlaid on cortical and subcortical brain templates. II: Scatter plots showing the relationships between ODI and [^11^C]UCB-J BP_ND_ for all regions surviving FDR correction, coloured by lobes and subcortex. Abbreviations: ODI = Orientation Dispersion Index. PSP-RS = Progressive Supranuclear Palsy-Richardson’s Syndrome; CBD = amyloid negative Corticobasal Syndrome.

**Supplementary Figure 1.**
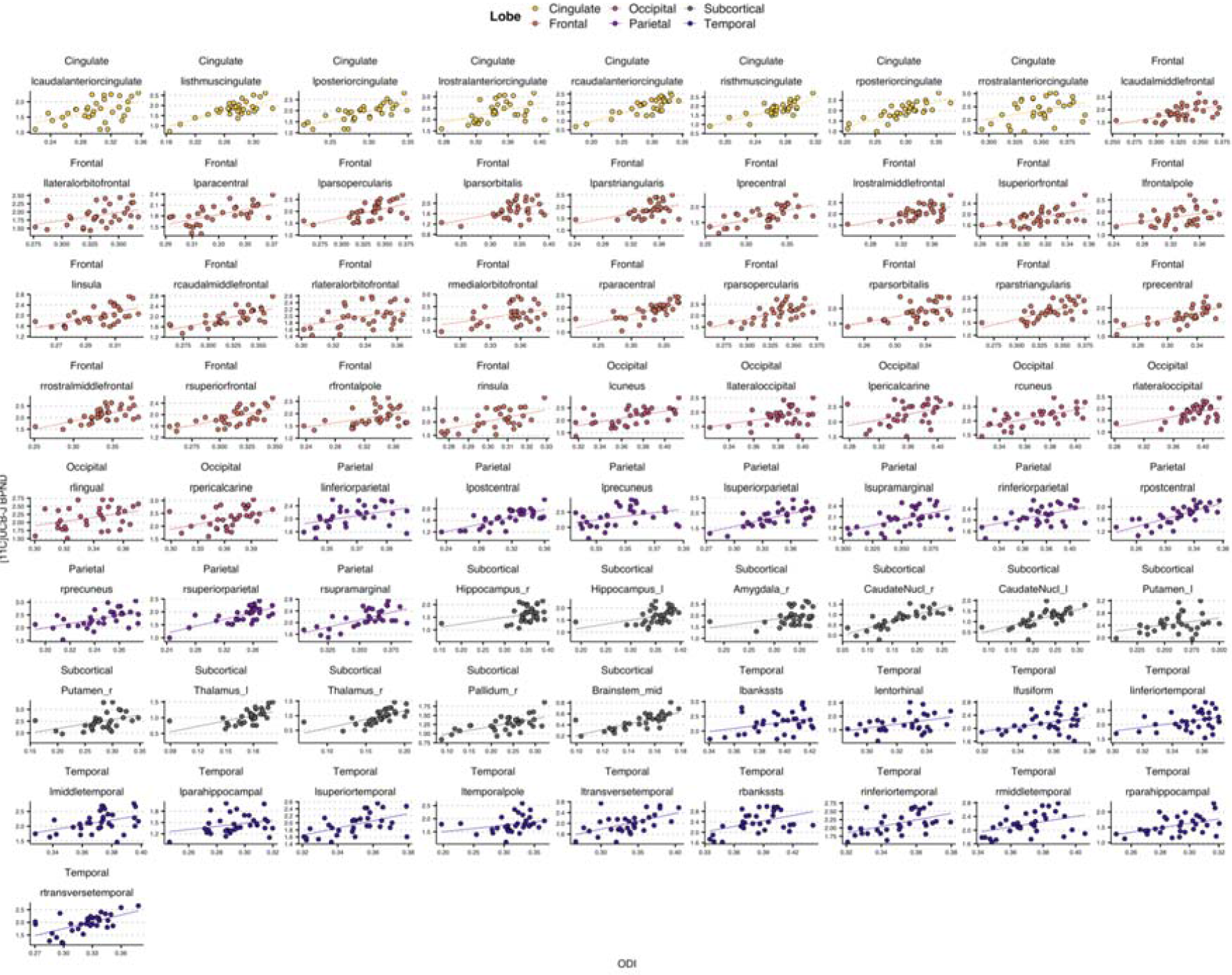
Significant local associations between cortical ODI and [^11^C]UCB-J BP_ND_ in PSP-RS and CBD patients (FDR p < 0.05, after adjusting for age and local GM atrophy). Abbreviation: ODI = Orientation Dispersion Index; FDR = False Discovery Rate; GM = Grey Matter.

## 4. DISCUSSION

This study tested the hypothesis that changes in synaptic density are closely linked to MRI measures of cortical microstructure, in the neurodegenerative tauopathies of CBD and PSP-RS. The main insights are that (i) in the neurodegenerative tauopathies of Corticobasal Degeneration (CBD) and Progressive Supranuclear Palsy (PSP-RS), widespread changes in grey matter dendritic complexity are observed even in areas without significant atrophy; and (ii) Orientation Dispersion Imaging (ODI), an MRI-based estimate of post-synaptic dendritic structure and complexity, correlates with the loss of presynaptic density estimated by the PET radioligand [^11^C]UCB-J; this effect is not a result of changes in grey matter atrophy. This extends previous investigations of ODI microstructural change in PSP-RS and CBS/CBD (Mitchell et al., 2019), demonstrating the *in vivo* coupling of dendritic complexity to presynaptic density, in line with preclinical models of tauopathies (Hoffmann et al., 2014; Rocher et al., 2010).

Grey matter atrophy within the motor cortex, as well as the basal ganglia and brainstem are common MRI findings in PSP-RS and CBD; the pattern of atrophy shown in figure 1 and 2 concords with previous studies (Jabbari et al., 2019). In this study however we show that, both within areas of the brain where there is atrophy, and in areas with absent atrophy, there is significant and more severe loss of dendritic complexity and, as previously shown (Holland et al., 2020) and replicated here, presynaptic density in the patient cohort. The widespread pattern of cortical ODI and presynaptic deficits in PSP-RS and CBD matches the expected pattern of tau pathology, both in the subcortical and cortical area (Dickson et al., 2011; Kovacs et al., 2020). Loss of dendritic complexity and presynaptic loss, in non-atrophied areas of the brain, for example the occipital lobes, potentially reflects early changes in synaptic function, in response to tau pathology (Kovacs et al., 2020), or toxic oligomers of tau, that later in the disease process progress to atrophy. Indeed, in preclinical models, pathological tau oligomers induce synaptic degeneration (Usenovic et al., 2015) and interfere with synaptic function and density (Yoshiyama et al., 2007), in the absence of neuronal loss.

The effect size for the comparison between ODI and [^11^C]UCB-J BP_ND_ in patients relative to controls, was stronger than for atrophy. Together, the larger extent of ODI reductions relative to grey matter atrophy indicate that NODDI can reveal new aspects of the cellular pathology of tauopathies, in keeping with the recent demonstration that changes in ODI parameters closely reflect complex histological changes (Grussu et al., 2017). Notwithstanding the cross-sectional design of this study, this observation highlights the early structural changes in disease pathogenesis that may underlie the emergence of cognitive and motor symptoms not attributable to atrophy. Indeed studies of cortical physiology in PSP-RS have illustrated abnormal electrophysiology in the absence of atrophy (Hughes et al., 2013; Sami et al., 2018). Our findings of reduced ODI beyond atrophy are in agreement with those published from the Alzheimer’s disease literature (Parker et al., 2018) which is a related tauopathy to PSP-RS and CBD, where there is a stronger relationship between cognitive function and synaptic density than with atrophy (Terry et al., 1991).

Our observation of a tight coupling between dendritic complexity (figure 3 and 4), as measured with ODI, and presynaptic density, as measured with [^11^C]UCB-J, in the patient cohort, echo preclinical findings in animal models of tauopathy (Harris and Kater, 1994), and post mortem studies (Bigio et al., 2001; Lipton et al., 2001). There are two potential explanation for this tight coupling: first, tau protein is enriched in axons and associated with axonal growth and transport; the synaptic toxicity associated with pathological tau is observed both in pre- and post-synaptic structures (see review by Mang & Mendelkow 2015 (Wang and Mandelkow, 2016)), leading to both a reduction in dendritic complexity and presynaptic density. Second, while a cross-sectional design precludes inferences of causality, alterations in synaptic function in CBD and PSP-RS (Holland et al., 2020) are expected to induce dendritic morphological alterations with early loss of dendritic spines and reduced dendritic branching. This has been illustrated in animal studies, where post-synaptic dendritic morphology correlates with the numbers of presynaptic vesicles and synaptic strength (Schikorski and Stevens, 1999). It is possible that primary dendritic degeneration in CBD and PSP-RS causes a reduction in the density of the presynaptic contacts on the distal dendrites. These alternative accounts are not mutually exclusive and could act in parallel to impair effective transneuronal connectivity, and subsequently motor and cognitive function.

The downstream effect of the above is a loss of functional connectivity. Indeed in PSP-RS there is reduced resting state connectivity between cortical and subcortical areas - reviewed in (Filippi et al., 2019). In CBD, the evidence is mixed as, thus far, functional MRI studies in this disease have not differentiated between amyloid positive and amyloid negative participants (also reviewed in (Filippi et al., 2019); in the former where Alzheimer’s pathology is the most likely finding at post mortem, functional connections are strengthened; however in the latter where a 4R tauopathy is likely, functional connections may become inefficient as seen in PSP-RS (Cope et al., 2018).

Synaptic pathology is a common feature in many neurodegenerative diseases (Clare et al., 2010; Herms and Dorostkar, 2016; Kweon et al., 2017). In the related tauopathy of Alzheimer’s disease, widespread reduction in synapses have been shown using [^11^C]UCB-J PET in patients (Mecca et al., 2020), as well as changes to dendritic complexity seen both in vivo as reduced ODI (Parker et al., 2018) and in animal models of Alzheimer’s disease (Dorostkar et al., 2015). Similar pre- and postsynaptic losses are seen in the substantia nigra of patients with Parkinson’s disease (Bellucci et al., 2016; Reeve et al., 2018), and within the hippocampi of patients with Lewy Body Dementia (Revuelta et al., 2008). The tight coupling of the presynaptic and post-synatpic compartments shown in our study, may therefore extend to pathologies other than primary 4R-tauopathies, where synaptic dysfunction is one of the earliest stages of disease, regardless of the culprit protein aggregate in question. If so, the opportunity to utilise both MRI and PET to understand disease pathogenesis would greatly facilitate cohort studies of individual phenotypic differences, or repeat testing in clinical trials monitoring. Furthermore, the *in vivo* observation of the tight coupling between the pre- and postsynaptic compartments, offer a new angle of interpretation for future studies utilising either of these methods in isolation; for example if cost, scalability and resources are limited for larger cohort studies, MRI measures of dendritic complexity can offer a close surrogate of presynaptic density, although we appreciate it is not a substitute given the significantly larger effect sizes seen with [^11^C]UCB-J (Figure 3C and 4C). This latter observation, which replicates our previous study in a smaller cohort of patients, may indicate a more severe toxic effect at the presynapse compared to dendritic pathology, although longitudinal studies are required to confirm this. However, it is important to note that ODI should not be taken as a direct representation of the post-synaptic density seen in histological studies. Rather, it provides complementary information to the PET data, with support from histological studies that have shown ODI to be a close surrogate (Grussu et al., 2017).

There are several strengths of this study. First, our NODDI resolution of 1.75mm was specifically optimised for the investigation of cortical microstructure, and is higher than previous NODDI studies in early onset AD (2.5 mm) (Parker et al., 2018), Parkinson’s disease (5 mm) (Kamagata et al., 2016) and PSP-RS (2.0 mm) (Mitchell et al., 2019). Second, we use amyloid imaging to exclude patients with CBS likely due to Alzheimer’s disease. This helps in reducing the potential underlying pathologies at play in the CBS cohort to 4R tauopathy, although we acknowledge other pathologies are possible but less likely (Alexander et al., 2014) – in this regard it is reassuring that the spatial patterns of reductions in ODI and [^11^C]UCB-J binding are not substantially different between the PSP-RS and CBD cohorts. Thirdly, the use of non-parametric permutation analyses also confer additional robustness to our statistical results, since this approach is not reliant on data distributions (Winkler et al., 2014).

We acknowledge the limitations of our study including the relatively small sample size in our CBD cohort. However, given the large effect sizes seen of ODI and [^11^C]UCB-J binding reduction in both our patient groups, our study was sufficiently powered to examine the relationships between both imaging markers. Secondly, we acknowledge that the MRI measure of ODI is not a measure of post-synaptic density but rather dendritic complexity; PET radioligands targeting the post-synaptic density may therefore provide further useful insights into post-synaptic pathology. Furthermore, although we illustrate a tight correlation between our two measures of synaptic health, we do not show causality in this cross-sectional study design, but are working towards this aim through our longitudinal studies. Lastly, we highlight the limitations of clinicopathological correlations, but take reassurance from previous validation studies illustrating a high consistency in the clinicopathological correlations in PSP-RS and in amyloid negative CBS.

Understanding the pathological processes that precede atrophy in neurodegeneration is key not only in expanding our knowledge of the pathophysiology of disease but also in informing the design of clinical trials both in terms of the imaging options for measuring disease-related changes and the optimal timing of intervention in the disease process. Our data implicate correlated changes in dendritic microstructure and synaptic density in patients with primary degenerative tauopathies including PSP-RS and CBD. Further cross-validation of ODI with [^11^C]UCB-J BP_ND_ may help further our understanding of the pathophysiology of neurodegeneration, applicable to future studies of early neurodegeneration with a safe and widely available MRI platform.

## Data Availability

The data that support the findings of this study are available from the corresponding author, upon reasonable request for academic (non-commercial) purposes, subject to restrictions required to preserve participant confidentiality.

## Abbreviations

PSP-RS: Progressive Supranuclear Palsy – Richardson’s Syndrome.
CBD: Corticobasal degeneration
PET: Positron Emission Tomography
NODDI: Neurite Orientation and Dispersion Imaging.

## ACKNOWLEDGMENTS

The authors thank the research participants and caregivers, the staff at the Wolfson Brain Imaging Centre, and at the Cambridge Centre for Parkinson-Plus. We thank UCB Pharma for providing the precursor for UCB-J synthesis. Infrastructure support was provided by the High Performance Hubs for Clinical Informatics (HPHI), funded by the MRC Research Infrastructure Award (MR/M009041/1).

